# The Effects of Intensive Antihypertensive Treatment Targets on Cerebral Blood Flow and Orthostatic Hypotension in Frail Older Adults

**DOI:** 10.1101/2023.10.05.23296632

**Authors:** Ralf W.J. Weijs, Bente M. de Roos, Dick H.J. Thijssen, Jurgen A.H.R. Claassen

**Affiliations:** Department of Medical BioSciences, Radboud university medical center, Nijmegen, The Netherlands; Department of Geriatric Medicine, Radboudumc Alzheimer Center, Donders Institute for Brain, Cognition and Behaviour, Radboud University Medical Center, Nijmegen, The Netherlands; Research Institute for Sport and Exercise Sciences, Liverpool John Moores University, Liverpool, United Kingdom; Department of Cardiovascular Sciences, University of Leicester, Leicester, United Kingdom

**Keywords:** frailty, geriatrics, longevity, orthostatic intolerance, primary health care, brain blood flow

## Abstract

**Background:** Guidelines recommend restrictive antihypertensive treatment (AHT) in hypertensive frail older adults, as intensive AHT is assumed to cause cerebral hypoperfusion and orthostatic hypotension (OH). However, studies directly examining these assumptions in older, frail individuals are lacking.

**Methods:** Fourteen frail hypertensive patients (six females; age 80.3±5.2 years; Clinical Frailty Scale 4-7; unattended SBP ≥150 mmHg) underwent measurements before and after a median of 7-weeks AHT (SBP target ≤140 mmHg). Transcranial Doppler measurements of middle cerebral artery velocity (MCAv), reflecting changes in cerebral blood flow (CBF), were combined with finger plethysmography recording of continuous BP. Transfer function analysis assessed cerebral autoregulation (CA). ANCOVA analyzed AHT-induced changes in CBF and CA, and evaluated non-inferiority of the relative change in CBF (margin: -10%; covariates: pre-AHT values and AHT-induced relative mean BP change). McNemar-tests analyzed whether the prevalence of (initial) OH, assessed by sit/supine-to-stand challenges, increased with AHT.

**Results:** Unattended mean arterial pressure decreased by 15 mmHg following AHT. Ten (71%) participants had good quality TCD assessments. Non-inferiority was confirmed for the relative change in MCAv (95%CI -2.7, 30.4). CA was normal and remained unchanged following AHT (*P*>0.05). None of the 14 participants had an increase in the prevalence of OH or initial OH (*P*≥0.655).

**Conclusions:** We found that AHT in frail, older patients does not reduce CBF, is not associated with impaired CA, and does not increase (initial) OH prevalence. These observations may open doors for more intensive AHT targets upon individualized evaluation and monitoring of hypertensive frail patients.

**Clinical Trial Registration:** ClinicalTrials.gov (NCT05529147) and EudraCT (2022-001283-10).

## Introduction

The global prevalence of hypertension has doubled in the past two decades,[1] which is relevant since hypertension represents a leading modifiable risk factor for burden of disease.[2] Large RCTs have established that anti-hypertensive treatment (AHT) is beneficial, with substantial (∼40%) risk reductions for cardiovascular events and mortality.[3, 4] Current treatment guidelines are moving towards lower SBP targets of 130-140 mmHg.[5, 6] Evidence supporting these guidelines mainly come from studies performed in non-frail individuals. For the rapidly increasing population of frail older patients, more conservative AHT is recommended.[5–8] These recommendations are based on the assumption that (intensive) AHT in frail older patients causes cerebral hypoperfusion or orthostatic hypotension (OH),[9, 10] leading to an increased risk for falls or dementia.[11] Importantly, the level of evidence supporting these recommendations is low, with few studies directly examining this relation.

The risk for cerebral hypoperfusion and OH with AHT in frail older individuals has long been linked to the widespread assumption that hypertension is a physiological, age-related adaptation to ensure sufficient CBF. However, this assumption has been questioned by recent studies. A recent meta-analysis found no negative effects of AHT on CBF in older hypertensive patients, including those with cognitive impairment.[12] Similarly, no evidence was found that AHT can impair cerebral autoregulation (CA) in older individuals.[13] Furthermore, in a large meta-analysis, intensive AHT did not increase the risk of OH.[14, 15] Together, these studies suggest that AHT does not cause cerebral hypoperfusion or OH in older, but mostly non-frail individuals. Evidence from studies in frail older adults however remains scarce. Therefore, the objective of this study was to directly examine the effects of intensive AHT, i.e. SBP ≤140 mmHg, on cerebral blood flow, cerebral autoregulation, and orthostatic hypotension, in a representative population of frail older adults.

## Methods

### Study design and participants

In this study, frail (Clinical Frailty Scale 4-7) older adults (age ≥70 years) with untreated or uncontrolled hypertension (unattended SBP ≥150 mmHg) were included for participation in this study. Detailed in- and exclusion criteria are listed in supplemental Table S1. Starting in September 2022, (records from) patients who visited the geriatric outpatient clinic between April 2022 and June 2023 were screened by a geriatrician for potential eligibility for participation in this explorative, single-arm intervention study. Potentially eligible patients received study information and, if they expressed interest in participation, were invited for a screening visit to confirm hypertension by an unattended BP measurement (at the clinic, or if preferred, at home). Before initiating or augmenting AHT, participants underwent baseline measurements in the research lab to assess cerebral hemodynamics, CA, and OH (lab visit 1). Subsequently, AHT was prescribed, and participants were visited two-weekly by a researcher to evaluate side effects or adverse events, and to perform home-based BP measurements. Once the treatment target (SBP ≤140 mmHg) was reached, lab measurements were repeated *in duplo* during follow-up on two separated days (lab visits 2 and 3 with 1-14 days in-between). A schematic overview of the study design with a participation flow-chart, is shown in Figure 1. The study was approved by the accredited local Medical Research Ethics Committee (METC Oost-Nederland; NL80929.091.22), registered with ClinicalTrials.gov (NCT05529147) and EudraCT (2022-001283-10), and conducted in accordance with the Declaration of Helsinki. All participants signed informed consent.

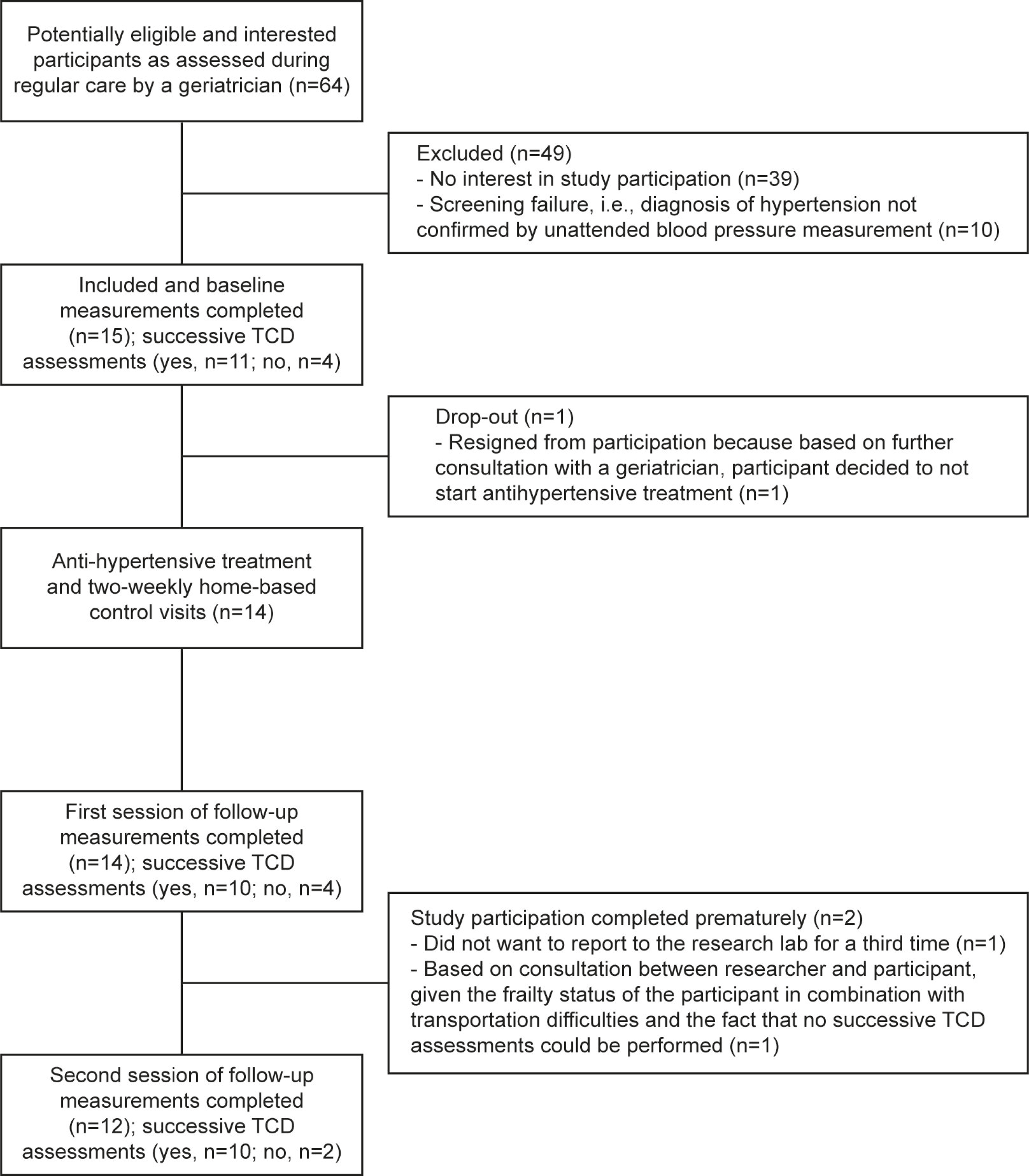

### Procedures

#### Participant characteristics

Relevant clinically collected data, e.g. during the geriatric outpatient visit, were derived from the electronic patient dossiers (EPD), including general demographics, Clinical Frailty Scale (CFS), Montreal Cognitive Assessment (MoCA) scores, the TOPICS-Short Form questionnaire (TOPICS-SF, a validated Dutch PROM), (Instrumental) Activities of Daily Living (IADL/ADL) scores, medical history and medication use. In case information for the MoCA and/or TOPICS-SF questionnaire was missing from the patient records, the MoCA and/or TOPICS-SF was completed during the baseline lab visit.

#### Blood pressure

BP measurements were performed using a Microlife WatchBP that automatically performs three assessments at the upper arm with 15-second rest intervals, and stores an averaged output. Unattended BP measurements were performed as part of screening and during the three lab visits. During these assessments, participants were sitting quietly for ≥5 min, alone, to reduce the risk of ‘white coat hypertension’ and to prevent interaction with others that could impact BP levels. The two-weekly home-based BP measurements were similarly performed, but in the presence of a researcher who avoided moving or talking.

#### Cerebral blood flow and autoregulation

During the three lab visits (one baseline and two follow-up), cerebrovascular measurements were performed under resting conditions with the patient seated on a chair for 5 min. Blood velocities in the left and right middle cerebral artery (MCA) were assessed using TCD (DWL Elektronische Systeme, Singen, Germany) by a trained ultrasonographer (RW). TCD is a non-invasive technique that applies ultrasound to track blood velocity changes in cerebral arteries, accessed through the transtemporal bone. Changes in mean bilateral blood velocity in the MCA (MCAv) represent changes in CBF under the assumption that the vessel diameter is constant, which has been confirmed under experimental conditions comparable to our study.[16] In addition, we measured beat-to-beat continuous BP using finger plethysmography (Finapres, Enschede, The Netherlands), three-lead electrocardiogram (Solar 8000M, GE Healthcare, Milwaukee, WI, USA) and end-tidal carbon dioxide (EtCO_2_) (BIOPAC Systems, Goleta, CA, USA). All data were recorded continuously using a data acquisition system (Acqknowledge; BIOPAC Systems, Goleta, CA, USA).

#### Orthostatic hypotension

To test for OH before and following AHT, participants performed a sit-to-stand and supine-to-stand postural change during the three lab visits. Following ≥5 min of seated/supine rest, patients were instructed to stand up to induce an orthostatic BP response, and to remain standing for 5.5 (sit-to-stand protocol) or 3.5 (supine-to-stand protocol) min while continuously recording heart rate (ECG) and beat-to-beat BP (finger plethysmography).

Sit-to-stand OH was defined as a drop of ≥15 mmHg in SBP or ≥7 in DBP after 1, 3 or 5 min of standing, while supine-to-stand OH was defined as a drop of ≥20 mmHg in SBP or ≥10 in DBP after 1 or 3 min of standing, relative to the resting value before standing up.[17] For both protocols, initial OH was defined in case the nadir, i.e. the lowest value upon standing up, was >40 mmHg in SBP or >20 in DBP lower compared to the resting value before standing up.[18] The supine-to-stand challenge more reliably tests for presence of OH and prognostic risk of falls compared to the sit-to-stand challenge.[19]

#### Anti-hypertensive treatment

In non-frail patients aged ≥65 years, European and Dutch guidelines for AHT advice a SBP target between 140-150 mmHg and, if well-tolerated, to consider a target between 130-139 mmHg.[5, 20] For frail older adults, guidelines let the treating physician decide the SBP target.[5, 20] For the purpose of this study, we have used the guidelines for non-frail patients aged ≥65 years, and used an unattended/home SBP target ≤140 mmHg. Individualized AHT was prescribed by a geriatrician (JC) after consideration of its safety for the participant, e.g. by taking renal and cardiovascular comorbidity and history into account. The medication classes used were angiotensin converting enzyme inhibitors, angiotensin II receptor blockers, calcium channel blockers, and/or thiazide diuretics. If indicated, betablockers and/or specific diuretics were additionally used for AHT. Rather than using a standard regimen, choices for drug class were based on comorbidity, co-pharmacy, and any allergies or contra-indications. In accordance with guidelines for older adults, the aim was to use the lowest dose for each class, with preference to add a second class rather than increase the dose of the first class when the BP target had not been reached.[20] In addition, the aim was to prescribe combination pills when possible.[20] Medication use was revised based on interim BP levels and possible side effects or adverse events monitored during the two-weekly home-based visits. To check medication adherence, participants were asked to record any deviations from the prescribed medication in a medication diary that was provided during the baseline lab visit.

### Data processing

SBP and DBP values from unattended/home BP measurements were used to calculate MAP values. AHT-induced BP change was expressed as the average of MAP values from *in duplo* unattended BP assessments during follow-up (lab visits 2 and 3) minus the MAP value from the unattended BP measurement during baseline (lab visit 1). Participants were numbered ordered from the smallest to the largest MAP reduction.

According to international guidelines,[21, 22] transfer function analyses of data recordings from TCD assessments during the 5-minute resting condition (including control parameters) were performed to derive the mean MCAv, MAP, and EtCO_2_, and to derive parameters of cerebral autoregulation (CA). CA parameters include transfer function gain, normalized gain, phase, and coherence over the low (0.07-0.20 Hz) and very low (0.02-0.07 Hz) frequency domains, where CA is most active. Cerebrovascular reactivity indices (CVRi) were calculated for each measurement (CVRi=MAP/MCAv). For each outcome parameter, single follow-up values were calculated by averaging the values from the *in duplo* follow-up assessments.

Continuous BP data recorded during the sit-to-stand and supine-to-stand challenges were automatically analyzed using MATLAB scripts to calculate mean SBP and DBP values over 30-second periods for seated/supine rest (i.e. 40-10 seconds before standing up), and for the first, third, and fifth minute after standing up (i.e. 45-75/105-135/285-315 seconds after standing up, respectively). In addition, the SBP and DBP nadir, i.e. the lowest value upon standing up, were identified. For each measurement, all automatically yielded outcomes were visually verified based on graphs created by MATLAB. Ultimately, absolute changes in BP values from seated or supine resting values were calculated for relevant timepoints.

### Statistical analyses

All statistical analyses were performed in IBM SPSS (version 27). Normal distribution of continuous variable data was checked visually. For descriptive data on participant characteristics at baseline, normally distributed data are presented as mean with standard deviation, and non-normally distributed data as median with interquartile range. Categorical data are presented as frequency number with percentage. Figures are used to show individual values from unattended/home BP measurements, resting hemodynamics and CA parameters, accompanied by means with 95% confidence intervals.

ANCOVA analyses were performed to compare baseline and follow-up values of resting hemodynamics and CA parameters with a random intercept and with baseline values and the medication induced change in MAP as covariates. The level of statistical significance was set at *P*<0.05. For our primary analysis, i.e. to test that AHT does not reduce CBF, these ANCOVA analyses were repeated for the relative change in MCAv from baseline to follow-up. Since we aimed to demonstrate absence of change, we adopted a non-inferiority design, with non-inferiority defined as the lower limit of the 95% confidence interval for the change in MCAv not exceeding the predefined margin of -10%. This margin was taken because 10% variation in CBF is expected under physiological conditions across a wide range of MAP, i.e. ∼50-150 mmHg.[13] AHT-induced changes in MCAv exceeding twice this variation, i.e. >20%, were carefully re-evaluated (e.g. to check for measurement error) to verify whether changes can be classified as non-physiological outliers. In case outliers were detected, ANCOVA analyses were repeated without individual data from these participants.

Individual BP data from sit-to-stand and supine-to-stand challenges are presented using spaghetti plots. The McNemar test was used to analyze whether paired proportions of initial OH and OH (i.e. yes/no based on both challenges) differed following AHT [23]. In addition, repeated measures ANOVA analyses were performed to examine whether SBP and DBP responses during sit-to-stand and supine-to-stand challenges were different between baseline and follow-up.

## Results

Fifteen hypertensive frail older adults were included in this study. One participant resigned from study participation before the initiation of AHT, leaving fourteen participants who completed study participation by undergoing baseline measurements and at least one session of follow-up measurements. Due to insufficient TCD signal quality, assessments of MCAv and CA were not performed in four (29%) of these participants. Details of these study procedures are presented in Figure 1, with participant characteristics and baseline medication use presented in Table 1 and 2, respectively.

**Table 1.** Participant characteristics.

|  | All (N=14) | With TCD (n=10) | Without TCD (n=4) |
| --- | --- | --- | --- |
| Age, years | 80.3±5.2 | 80.1±6.1 | 80.1±2.1 |
| Female sex, n (%) | 6 (43) | 2 (20) | 4 (100) |
| Unattended BP during screening |  |  |  |
| SBP, mmHg | 164±10 | 163±9 | 167±13 |
| DBP, mmHg | 87±11 | 90±11 | 78±6 |
| SBP 150-159 mmHg, n (%) | 6 | 4 | 2 |
| SBP ≥160 mmHg, n (%) | 8 | 6 | 2 |
| Marital status |  |  |  |
| Married, n (%) | 7 (50) | 6 (60) | 1 (25) |
| Widow(er), n (%) | 6 (43) | 3 (30) | 3 (75) |
| Divorced, n (%) | 1 (7) | 1 (10) | 0 (0) |
| Living situation |  |  |  |
| Independently together, n (%) | 7 (50) | 6 (60) | 1 (25) |
| Independently alone, n (%) | 6 (43) | 3 (30) | 3 (75) |
| Retirement home, n (%) | 1 (7) | 1 (10) | 0 (0) |
| Education level [33] |  |  |  |
| Low, n (%) | 7 (50) | 4 (40) | 3 (75) |
| Middle, n (%) | 5 (36) | 4 (40) | 1 (25) |
| High, n (%) | 2 (14) | 2 (20) | 0 (0) |
| Smoking status |  |  |  |
| Never smoked, n (%) | 7 (50) | 4 (40) | 3 (75) |
| Former smoker, n (%) | 5 (36) | 4 (40) | 1 (25) |
| Current smoker, n (%) | 2 (14) | 2 (20) | 0 (0) |
| Height, cm | 169±10 | 172±9 | 161±9 |
| Weight, kg | 79.7±18.4 | 84.0±19.4 | 69.0±11.0 |
| Body mass index, kg/m<sup>2</sup> | 26.6 (24.5-29.2) | 26.6 (24.5-30.1) | 26.7±3.1 |
| Estimated GFR, ml/min/1.73m<sup>2</sup> | 70.5±16.0 | 70.7±16.0 | 70.3±16.2 |
| Arterial sodium, mmol/l | 140±2 | 140±2 | 141±2 |
| Arterial potassium, mmol/l | 4.1±0.3 | 4.1±0.3 | 4.2±0.3 |
| CFS 4:5:6:7, n (%) | 7:4:3:0<br>(50:29:21:0) | 5:3:2:0<br>(50:30:20:0) | 2:1:1:0<br>(50:25:25:0) |
| Subjective health, score (1-10) | 6.6±1.0 | 6.8±1.0 | 6.3±1.0 |
| Subjective quality of life, score (1-10) | 7.2±1.5<sup>a</sup> | 7.3±1.6<sup>a</sup> | 7.0±1.4 |
| ADL dependency score 0:1:2:3<sup>b</sup>, n (%) | 7:5:2:0 | 6:3:1:0 | 1:2:1:0 |
| **Instrumental ADL dependency score 0:1:2:3<sup>b</sup>, n (%)** | (50:36:14:0)<br>6:1:7:0<br>(43:7:50:0) | (60:30:10:0)<br>5:1:4:0<br>(50:10:40:0) | (25:50:25:0)<br>1:0:3:0<br>(25:0:75:0) |
| **MoCA, total score (0-30)** | 19.8±3.8 | 20.7±3.1 | 17.5±5.1 |
| **MoCA, MIS score (0-15)** | 8.1±4.4 | 7.8±3.5 | 9.0±6.7 |
| **Subjective memory complaints, n (%)** | 11 (79) | 9 (90) | 2 (50) |
| **Clinical cognitive decline, n (%)** | 10 (71) | 7 (70) | 3 (75) |
| **Mild cognitive impairment, n (%)** | 8 (57) | 5 (50) | 3 (75) |
| **Dementia, n (%)** | 2 (14) | 2 (20) | 0 (0) |
| **Anxiety disorders, n (%)** | 1 (7) | 0 (0) | 1 (25) |
| **Depression, n (%)** | 4 (29) | 2 (20) | 2 (50) |
| **Asperger's syndrome, n (%)** | 1 (7) | 1 (10) | 0 (0) |
| **Angina, coronary heart disease or myocardial infarction, n (%)** | 5 (36) | 3 (30) | 2 (50) |
| **Atrial fibrillation/flutter, n (%)** | 3 (21) | 2 (20) | 1 (25) |
| **Cardiac valve disease, n (%)** | 4 (29) | 2 (20) | 2 (50) |
| **Heart failure, n (%)** | 1 (7) | 0 (0) | 1 (25) |
| **Hypertension, n (%)** | 14 (100) | 10 (100) | 4 (100) |
| **Peripheral vascular disease, n (%)** | 2 (14) | 2 (20) | 0 (0) |
| **Stroke/TIA, n (%)** | 1 (7) | 0 (0) | 1 (25) |
| **Small vessel disease, n (%)** | 1 (7) | 1 (10) | 0 (0) |
| **Chronic kidney disease (estimated GFR<60)** | 3 (21) | 2 (20) | 1 (25) |
| **Diabetes** | 3 (21) | 1 (10) | 2 (50) |
| **Arthritis/arthrosis/osteoporosis** | 3 (21) | 2 (20) | 1 (25) |
| **Cancer within past 5 years** | 2 (14) | 1 (10) | 1 (25) |
| **Asthma / COPD** | 1 (7) | 1 (10) | 0 (0) |
| **Visual/auditory impairment, n (%)** | 8 (57) | 6 (60) | 2 (50) |
| **Polyneuropathy, n (%)** | 3 (21) | 2 (20) | 1 (25) |
| **Balance problems, n (%)** | 9 (64) | 7 (70) | 2 (50) |
| **Dizziness, n (%)** | 8 (57) | 5 (50) | 3 (75) |
| **Fall within past year, n (%)** | 7 (50) | 5 (50) | 2 (50) |

<sup>a</sup>1 missing value. <sup>b</sup>Score categories: 0=fully independent, 1=moderately impaired, 2=severely impaired, 3=fully dependent [34].
Abbreviations: ADL, activity of daily living; BP, blood pressure; DBP, diastolic blood pressure; COPD, chronic obstructive pulmonary disease; GFR, glomerular filtration rate; SBP, systolic blood pressure; TCD, transcranial Doppler; TIA, transient ischemic attack.

**Table 2.** Medication use at baseline.

| **Medication use, n (%)** | **All (N=14)** | **With TCD (n=10)** | **Without TCD (n=4)** |
| --- | --- | --- | --- |
| **Polypharmacy (use of ≥5 prescribed medications)** | 11 (79) | 7 (70) | 4 (100) |
| **Cardiovascular medication** |  |  |  |
| **Antihypertensives** | 12 (86) | 8 (80) | 4 (100) |
| **Betablocker(s)** | 6 (43) | 3 (30) | 3 (75) |
| **ACE inhibitor(s)** | 5 (36) | 3 (30) | 2 (50) |
| **ARB** | 4 (29) | 3 (30) | 1 (25) |
| **CCB** | 4 (29) | 3 (30) | 1 (25) |
| Thiazide diuretic(s) | 2 (14) | 1 (10) | 1 (25) |
| Spironolactone | 2 (14) | 0 (0) | 2 (50) |
| Statins | 5 (36) | 2 (20) | 3 (75) |
| Loop diuretic(s) | 1 (7) | 1 (10) | 0 (0) |
| Digoxin | 1 (7) | 0 (0) | 1 (10) |
| Nitrates | 2 (14) | 2 (20) | 0 (0) |
| Salicylates | 4 (29) | 3 (30) | 1 (10) |
| Anti-coagulants | 4 (29) | 3 (30) | 1 (10) |
| Platelet-inhibitor(s) | 1 (7) | 1 (10) | 0 (0) |
| DOAC | 3 (21) | 2 (20) | 1 (10) |
| Diabetes medication |  |  |  |
| Biguanides | 2 (14) | 0 (0) | 0 (0) |
| Psychotropic medication |  |  |  |
| Antidepressants | 4 (29) | 2 (20) | 2 (50) |
| Tri-/tetracyclic antidepressant(s) | 3 (21) | 1 (10) | 2 (50) |
| SSRI | 1 (7) | 1 (10) | 0 (0) |
| Benzodiazepines | 1 (7) | 0 (0) | 1 (25) |
| Antiepileptic | 1 (7) | 1 (10) | 0 (0) |
| Cannabidiol oil | 1 (7) | 1 (10) | 0 (0) |
| Urological medication use |  |  |  |
| Alfa blocker | 1 (7) | 1 (10) | 0 (0) |
| 5-alpha-reductase inhibitors | 1 (7) | 1 (10) | 0 (0) |
| Spasmolytics | 3 (21) | 3 (30) | 0 (0) |
| Other medication use |  |  |  |
| Bisphosphonates | 1 (7) | 1 (10) | 0 (0) |
| Proton pump inhibitors | 6 (43) | 3 (30) | 3 (75) |
| Vitamin/mineral supplements | 8 (57) | 7 (70) | 1 (25) |
| Inhalational parasympatholytics | 1 (7) | 1 (10) | 0 (0) |
| Laxatives | 4 (29) | 3 (30) | 1 (25) |
| Antispasmodics | 1 (7) | 0 (0) | 1 (25) |

### Medication induced blood pressure changes

In thirteen participants (93%), AHT successfully reduced SBP to the treatment target of ≤140 mmHg (supplemental Figure S1A) across 6±3 weeks of AHT. In one patient (7%), home and unattended SBP remained above target (i.e. 162 and 154 mmHg, respectively; supplemental Figure S1A and Figure 2) despite 22 weeks of AHT, indicating uncontrolled hypertension. On average, unattended MAP was reduced across follow-up by 15±12 mmHg (Figure 2). All participants were included in the analyses. For an overview of medication used for AHT, see supplementary Table S2.

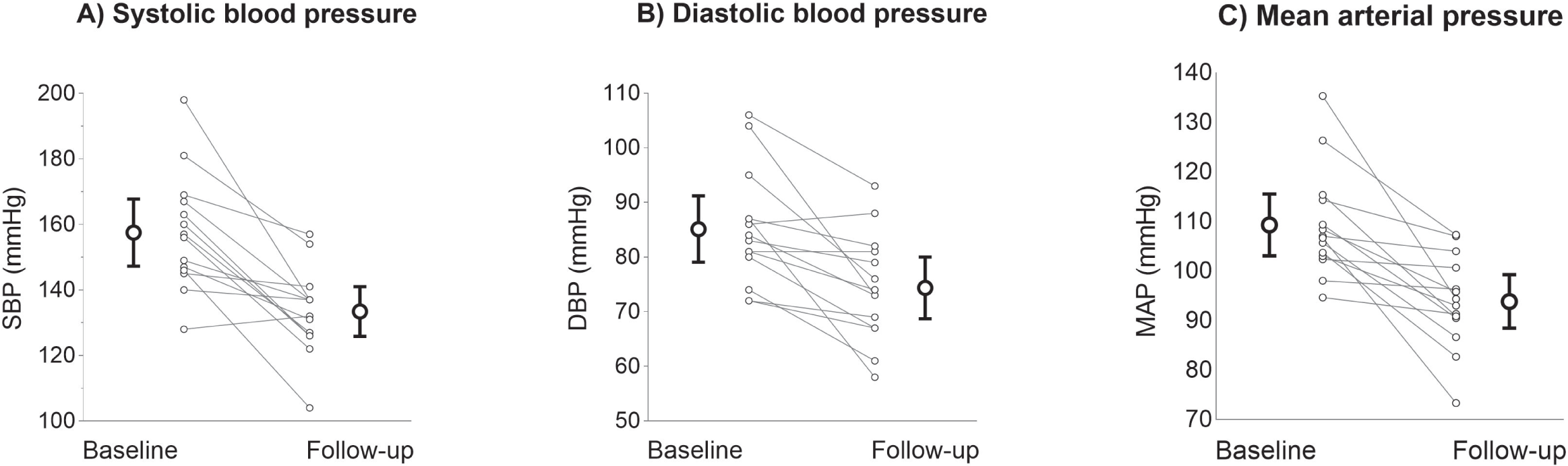

### Cerebral artery blood velocity and cerebral autoregulation

ANCOVA analyses revealed no absolute change in MCAv across follow-up (Figure 3A), whereas CVRi was reduced significantly (Figure 3B). Regarding our primary outcome analysis, the change in MCAv following AHT (mean=13.9%; 95%CI=-2.7, 30.4) did not cross the non-inferiority margin of -10% (Figure 3C). At individual level, one participant exceeded the non-inferiority margin for MCAv (i.e. -15%). This participant also demonstrated the largest AHT-induced MAP reduction (i.e. -25 mmHg). We identified one non-physiological outlier, as insonation depths during TCD assessments at baseline and during follow-up were not comparable. Repeating the ANCOVA analysis without this individual reinforced non-inferiority (10.7%; 95%CI = -7.4, 28.7).

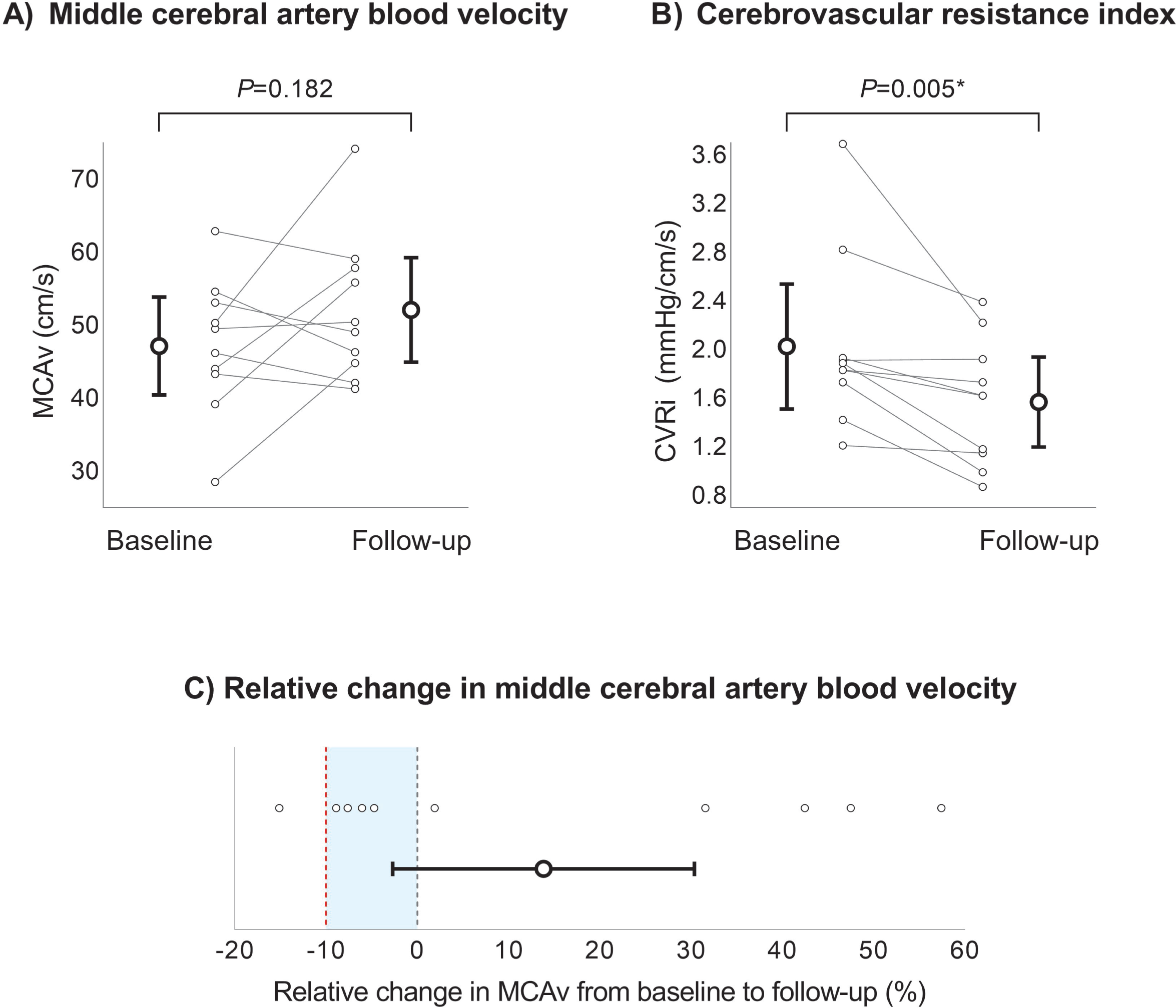

For CA, a statistically significant change was found only in one of eight parameters, i.e. transfer function gain in the low frequency domain (supplemental Figure S2). However, all other CA parameters over the low (Supplemental Figure S2) and very low frequency range (supplemental Figure S3) remained unchanged during AHT. Repeating these analyses without the statistical outlier did not alter the outcomes (supplemental Table S3).

### Orthostatic tolerance

All participants performed a sit-to-stand challenge at baseline and at least one sit-to-stand challenge at follow-up (Figure 5A-B). The supine-to-stand challenge was performed at baseline and at least once during follow-up by 12 (86%) participants (Figure 5C-D). During the sit-to-stand challenge, three individuals (21%) met the criteria for OH at baseline for SBP, which remained present for one (7%) during follow-up. This participant (#7) also met criteria for OH during the supine-to-stand challenge at baseline and during follow-up. None of the others met criteria for OH following AHT based on the sit-to-stand challenge. During the supine-to-stand challenge at baseline, only participant #7 met the criteria for OH. Two participants (#5 and #13) developed OH following AHT during the supine-to-stand challenge, with OH being symptomatic (dizziness) in one of them. Based on both challenges, prevalence of OH and initial OH did not change after AHT (OH: 21% to 21%, *P*=1.000; initial OH: 36% to 43%, *P*=0.655). Following AHT, absolute changes in SBP and DBP during sit-to-stand and supine-to-stand challenges did not differ from those at baseline (sit-to-stand: SBP, *P*=0.846 and DBP, *P*=0.898; supine-to-stand: SBP, *P*=0.462 and DBP, *P*=0.823).

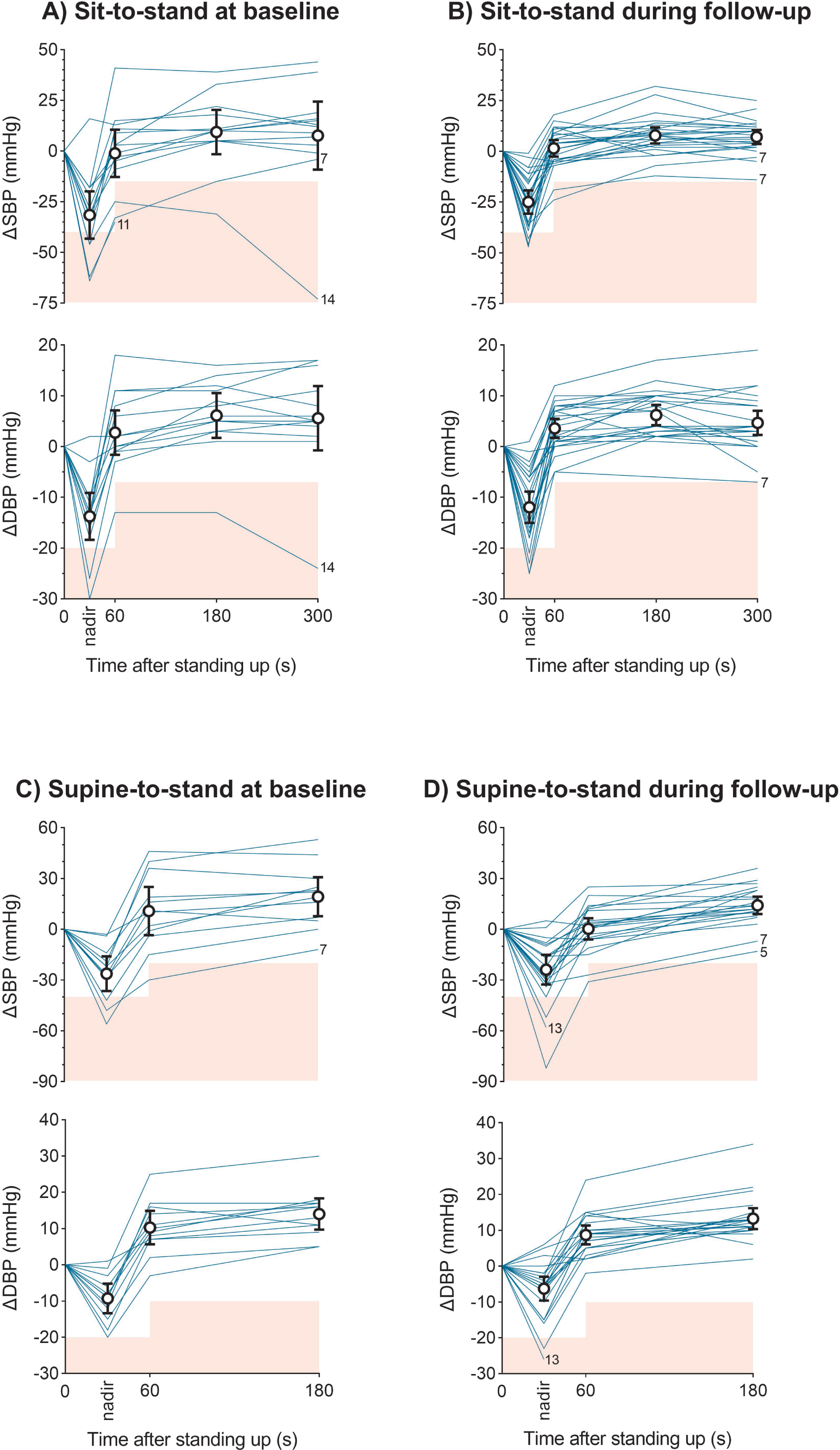

## Discussion

The aim of this study was to explore the effect of intensive AHT in frail older adults on cerebral blood flow, cerebral autoregulation, and prevalence of orthostatic hypotension. AHT successfully lowered unattended MAP by 15 mmHg, and met the SBP target of ≤140 mmHg in 13 out of 14 participants. Confirming our non-inferiority hypothesis, we found that AHT did not reduce CBF in older, frail individuals. In line with this result, CA remained within normal ranges following AHT. Regarding postural hypotension, we found no change in the prevalence of OH between baseline and AHT, and continuously measured BP responses during sit-to-stand or supine-to-stand postural changes were not affected by AHT. Altogether, our observations indicate that AHT with a treatment target of SBP ≤140 mmHg in this group of frail, older individuals did not cause cerebral hypoperfusion, impairment in CA, or higher prevalence of OH, which may have clinical relevance in the prescription of AHT in this population.

Confirming our hypothesis, intensive AHT in older, frail individuals is not accompanied by a reduction in CBF. This observation is in line with previous findings in non-frail older adults.[12, 24] For example, a recent meta-analysis, which includes the study by Lipsitz *et al.*,[24] reported stable CBF following AHT-induced BP reductions in adults aged ≥50 years, while subgroup analysis even revealed increases in CBF in those aged >70 years.[12] In addition, none of the six studies included in this meta-analysis that examined patients with mild cognitive impairment or dementia found AHT-induced changes in CBF. This is relevant, because cognitive decline is associated with (higher risk of) disturbed cerebrovascular physiology and, in the context of our study, is considered a feature of frailty. It should be stressed, however, that not every individual with cognitive decline is frail, and that not every frail individual experiences cognitive decline. As frailty represents a complex multifactorial concept, including contributing factors such as older age and cognitive decline that have not been associated with AHT-induced CBF reduction, our study adds the novel information that CBF is not reduced following AHT in a well-defined representative group of older, frail individuals.

It has been suggested that age-related hypertension represents a physiological adaptation, required to maintain CBF as older age leads to irreversible cerebrovascular stiffening.[25, 26] However, this theory has been challenged recently by evidence indicating that the cerebrovasculature has the ability to adapt.[12] Our observation that CBF is not altered following AHT supports this notion, although we explored the effects of AHT on cerebrovascular parameters across a relatively short period. Long-term studies with repeated follow-up measurements are required to investigate whether these beneficial vascular adaptations persist to maintain sufficient CBF. This is especially relevant, as AHT primarily aims to provoke extended healthy longevity by reducing risk of morbidity and mortality by cardio- and cerebrovascular disease. Moreover, (intensive)[3] AHT reduces the risk of cognitive decline and dementia.[27] Given recent observations that reductions in CBF are associated with cognitive decline,[28, 29] the apparent effect of AHT to increase CBF may contribute to the clinical potential of AHT for the prevention of dementia in frail individuals.

Previous work had suggested that AHT-induced CBF reductions in non-frail hypertensives may be related to a rightward shift in the CA curve.[30] Consequently, AHT would lead to impaired CA in case decreased BP levels fall below the lower limit of the rightward-shifted CA curve. Therefore, we examined AHT-induced changes in CA, and observed an isolated increase in transfer function gain over the low frequency domain. This may be related to the change in cerebrovascular resistance following AHT.[31] Compared with normative values in healthy individuals,[21] our population indeed demonstrated a relatively low gain before initiation of AHT, while following AHT, values approximated normative values, consistent with observations in earlier studies.[31] While an increase in transfer function gain is frequently interpreted as impaired CA, it is important to combine the results of all transfer function parameters. The combined results of phase and gain in the frequency domains we studied indicate normal and unchanged CA.

To further investigate the potential risks associated with AHT in frail older adults, we explored the prevalence of both OH and initial OH, which, from a clinical perspective, represent a major concern as it causes dizziness and is associated with increased risk of falls. However, our results demonstrate no change in the prevalence of OH nor of initial OH. In addition to examining OH as a binary outcome using strict cut-off values, we also examined the absolute changes in BP, which reinforces our initial observation that AHT does not alter BP responses to orthostatic challenges in frail, older hypertensives. This is in line with previous evidence, primarily in non-frail individuals, concluding that intensive AHT reduces the risk of OH,[14] while deprescribing AHT may increase the risk of OH.[32] Altogether, hypertension seems an important risk factor for OH, with lowering BP to ‘normal’ levels unlikely being a risk factor for OH, even in older, frail individuals.

### Strengths and limitations

Strengths of our study include the prospective, controlled design, and the inclusion of a well-defined and representative older population of frail individuals, recruited form a geriatric outpatient clinic. Strengths also include the comprehensive, *in duplo* evaluation of main outcome parameters. However, some limitations must be considered. First, our statistical analyses regarding the prevalence of (initial) OH may be underpowered. However, our comprehensive protocol (including two orthostatic challenges, performed *in duplo*) and BP analysis on a continuous scale, strongly support our conclusion that AHT does not increase OH prevalence. Second, a potential limitation is that the vasodilator effects of AHT may have caused an increase in MCA diameter during follow-up. Under this assumption, we may have underestimated the true CBF following AHT. At least, this supports our conclusion that AHT does not cause cerebral hypoperfusion in frail older adults.

### Perspectives

Our observations suggest that successful blood pressure lowering following intensive AHT in frail older adults does not lead to cerebral hypoperfusion, impaired cerebral autoregulation, and/or increased prevalence of orthostatic intolerance. These observations argue against the assumption that AHT in frail individuals must be prevented because of the risk for cerebral hypoperfusion or orthostatic hypotension. Our results, therefore, strongly support future studies to examine the longer-term effects of intensive AHT in frail older adults, which is further supported by the clinical benefits of intensive AHT in the prevention of cardiovascular disease and dementia. Nonetheless, it remains important that AHT prescription is based on individualized comprehensive evaluation, especially pertaining to the heterogeneous responses observed in our study. As our study was not designed to specifically address mechanisms, we support future studies to better understand the (short- and long-term) potential risks and benefits of intensive AHT in frail older individuals, and to identify mechanisms explaining the preservation of cerebral perfusion upon AHT in this frail, older population.

## Data Availability

The data of this study are available upon reasonable request from the corresponding author.

## Nonstandard Abbreviations and Acronyms

AHT: Antihypertensive treatment
CBF: Cerebral blood flow
CA: Cerebral autoregulation
MCAv: Mean bilateral middle cerebral artery blood velocity
OH: Orthostatic hypotension
TCD: Transcranial Doppler

## Novelty and Relevance

### What is New?

- This study explored whether intensive antihypertensive treatment (AHT) causes cerebral hypoperfusion and/or orthostatic hypotension (OH) in older, frail hypertensives.

### What is Relevant?

- Intensive AHT does not lead to reductions in CBF, impaired CA, or increased prevalence of OH.
- Our findings suggest that intensive blood pressure management in older, frail adults is safer than was hitherto assumed, and can be considered for prevention of cardiovascular events and cognitive decline.

### Clinical/Pathophysiological Implications?

- Side effects of AHT in frail older adults may be overstated in literature and guidelines.
- With personalized evaluation and monitoring, a SBP target <140 mmHg can be achieved in older, frail populations without causing cerebral hypoperfusion or orthostatic hypotension.

## Acknowledgements

We would like to thank everyone who contributed to conducting this study. We are grateful to all study participants and their caregivers for their time and effort.

